# Antibiotic Resistance in Wastewater Treatment Plants and Transmission Risks for Employees and Residents: The Concept of the AWARE Study

**DOI:** 10.1101/2021.02.03.21250668

**Authors:** Laura Wengenroth, Fanny Berglund, Hetty Blaak, Mariana Carmen Chifiriuc, Carl-Fredrik Flach, Gratiela Gradisteanu Pircalabioru, D. G. Joakim Larsson, Luminita Marutescu, Mark van Passel, Marcela Popa, Katja Radon, Ana Maria de Roda Husman, Daloha Rodríguez-Molina, Tobias Weinmann, Andreas Wieser, Heike Schmitt

## Abstract

**Background:** Antibiotic resistance has become a serious global health threat. Wastewater treatment plants (WWTPs) may become unintentional collection points for bacteria resistant to antimicrobials. Little is known about the transmission of antibiotic resistance from wastewater treatment plants to humans, most importantly to WWTP workers and residents living in the vicinity. We aim to deliver precise information about the methods used in the AWARE (Antibiotic Resistance in Wastewater: Transmission Risks for Employees and Residents around Waste Water Treatment Plants) study.

**Methods/Design:** Within the AWARE study, we gather data on the prevalence of two antibiotic resistance phenotypes, ESBL-producing *E*.*coli* (ESBL-EC) and carbapenemase-producing Enterobacteriaceae (CPE) as well as on their corresponding antibiotic resistance genes (ARGs) isolated from air, water, and sewage samples taken from inside and outside of different WWTPs in Germany, Netherlands and Romania. Additionally, we analyse stool samples of WWTP workers, nearby residents and members of a comparison group living ≥1,000 m away from the closest WWTP.

**Discussion:** The study results will enable the assessment of the potential health impact of exposure to ESBL-EC, CPE and ARGs in and around WWTPs. Quantifying the contribution of different wastewater treatment processes to the ESBL-EC, CPE and ARGs removal efficiency will provide us with evidence-based support for possible mitigation strategies.

## Background

Antibiotic resistance has become a serious global health threat. As bacteria and certain genetic traits often move between humans, animals and the environment, a one health approach that considers these interactions is needed to efficently address this growing problem. The role of the environment in the emergence and dissemination of antibiotic resistance has become more and more acknowledged (Bengtsson-Palme et al. 2018;Berendonk et al. 2015; Martinez et al. 2009). Still, little is known about the transmission dynamics of antibiotic resistance determinants from water, air and soil and their risks for humans in direct contact with these matrices (Larsson et al. 2018). A key to determining human health impact lies in the application of epidemiological investigations, in which carriage of antibiotic resistant bacteria (ARB) in people exposed to a specific transmission route is tested in comparison to unexposed or less exposed controls. Such studies have been carried out in travellers (Paltansing et al. 2013) and in agricultural settings (Carrel et al. 2014; Graveland et al. 2010), but other environmental exposure routes, such as via water have rarely been studied (Huijbers et al. 2015; Leonard et al. 2018; Rodriguez-Molina et al. 2019; Soraas et al. 2013; Wuijts et al. 2017).

Wastewaters from agriculture, industry, hospitals, and households are collected together at wastewater treatment plants (WWTPs), making them unintentional collection points for antimicrobials and ARB. Wastewater typically harbours a mix of residual antibiotics, and other agents that are known to co-select for antibiotic resistance (Pal et al. 2015; Pal et al. 2014), which provides opportunities for selection of ARBs and hence risks for evolution and transmission of resistance. Selection pressures, together with a high density and diversity of pathogens and environmental bacteria carrying various antibiotic resistance factors provide a milieu where new forms of resistance may emerge (Finley et al. 2013; Gaze et al. 2013). From mining of metagenomics data, we know that emergence of new antibiotic resistance genes (ARG) occurs (Berglund et al. 2017; Boulund et al. 2012). Additionally, resistant bacteria already present in human feces can pass WWTPs. E.g. ESBL-producing *E*.*coli* (ESBL-EC) have been detected in the influent and effluent of WWTPs and the receiving surface waters (Brechet et al. 2014). It is known that human infections with ESBL-EC or carbapenem-resistant Enterobacteriaceae (CPE) are associated with increased mortality rates, time to effective therapy, length of hospital stay and overall healthcare costs (Wilson et al., 2018).

WWTPs are in general not developed to remove either of these (or any) resistant bacteria. Studies indicate that even though a significant reduction occurs through various treatment processes, significant amounts of antimicrobials, ARB and ARGs are still shed into environmental reservoirs, including rivers and recreational water (Pruden et al. 2013). While the efficiency of conventional treatment technologies greatly differs between types of WWTPs, the role of specific treatment technologies in removal of antimicrobials, ARB and ARGs remains poorly described (Pallares-Vega et al. 2019; Rizzo et al. 2013).

Workers at WWTPs are potentially exposed to wastewaters carrying ARB and ARGs and aerosolised ARB and ARGs through different transmission routes: inhalation, dermal contact, and ingestion. Airborne bacteria have indeed been detected in WWTPs (Cyprowski et al. 2018; Xu et al. 2020; Yang et al. 2019), including *Enterobacteriaceae* and fecal coliforms (Heinonen-Tanski et al. 2009; Xu et al. 2018), and an increased prevalence of gastrointestinal and respiratory diseases was reported in WWTP workers, suspected to be linked to microbial exposures (Thorn and Beijer 2004). Although few studies so far addressed specific pathogens in WWTP workers, one has found an elevated carriage of *Tropheryma whipplei* (Schöniger-Hekele et al. 2007). Additionally, a higher seroprevalence of IgG against *Helicobacter pylori* was observed among sewage workers (Van Hooste et al. 2010). Hepatitis A virus, hepatitis E virus and positive stool PCR tests for *Leptospira spirochete* (Albatanony and El-Shafie 2011) were also described. However, the carriage of ARB and ARGs in WWTP workers is yet unknown.

Furthermore, WWTPs are often located in urban settings in close proximity to residents. As bacteria can be traced back up to 150 meters away from animal farms (Gilchrist et al. 2007), neighbouring residents might also face a risk of exposure to aerosolized wastewater. WWTPs, their workers and nearby residents therefore could represent an ideal - but yet unstudied - test case to investigate whether transmission via (waste)water actually impacts ARB and ARGs carriage.

## Aims of the AWARE study

Within the AWARE study (“Antibiotic Resistance in Wastewater: Transmission Risks for Employees and Residents around Waste Water Treatment Plants”), we gather data on two antibiotic resistance phenotypes i.e ESBL-producing *E*.*coli* (ESBL-EC) and carbapenem-resistant *Enterobacteriaceae* (CPE) and on ARGs prevalence from analysis of air, water, sewage and stool samples taken from inside and outside of different WWTPs in Germany, Netherlands and Romania. The AWARE study specifically aims:

- to study carriage rates of ESBL-EC, CPE and of a range of clinically relevant ARGs in WWTP workers and nearby residents (living within ≤300 m vicinity of a WWTP) compared to a comparison group (living 1,000m away from the closest WWTP).
- to study waterborne and airborne exposure to ESBL-EC, CPE and of a range of clinically relevant ARGs in WWTP workers through ingestion and inhalation,
- to assess the efficiency of different WWTP treatment technologies in diminishing ESBL-EC, CPE and a range of clinically relevant ARGs,
- to investigate selection and emergence of ESBL-EC, CPE and a range of clinically relevant ARGs in WWTPs through studying relative changes in resistance genes and exploring putative novel resistance genes from metagenomics data.

Our overall aim with this methodological publication is to deliver precise information about the methods used in the AWARE project, including selection of participants, sample taking, creation of the questionnaire and pilot study. Further, we will discuss possible strengths and limitations of our study design.

## Methods/Design

### Study design

The AWARE study is a multicentre, cross-sectional study investigating the prevalence of ESBL-EC, CPE and ARGs in WWTP workers, residents living within ≤300 m vicinity of a WWTP (residents) and a comparison group living >1,000 m away from the closest WWTP (comparison group). The field phase is carried out in Germany (DE), the Netherlands (NL) and Romania (RO).

### Study population

We aim to include 450 WWTP workers (150 per country). In order to compare carriage of ESBL-EC, CPE and ARGs we aim to include 800 nearby residents (400 in DE, 400 in RO) living in <300 m vicinity of a WWTP (residents). Further, we aim to include 1,200 residents (400 in DE, 400 in RO, 400 in NL) living >1,000 m away from the closest WWTP (comparison group). Assuming an average ESBL-EC prevalence of 8% in the general population, this would allow us detecting a minimum odds ratio (OR) of 1.7 with power 80% in workers and nearby residents on a 5% significance level.

In order to be included in the study, participants have to be within the age range of 16 to 67 years. All participants who have worked at a slaughterhouse or a farm during 12 months prior to study are excluded because contact with farm animals and working at slaughterhouses can be risk factors for ESBL carriage (Dohmen et al. 2017).

### Recruitment process

The recruitment process for WWTP workers, residents living within ≤300 m of a WWTP and the comparison group consisting of residents living >1,000 m away from the closest WWTP is underlying local regulations and thus differs between DE, NL and RO (Table 1). However, to control for seasonal variation of ESBL-EC, CPE and ARGs, we aim to take all samples (water, air, stool) from the surroundings of each WWTP within eight weeks.

**Table 1.**
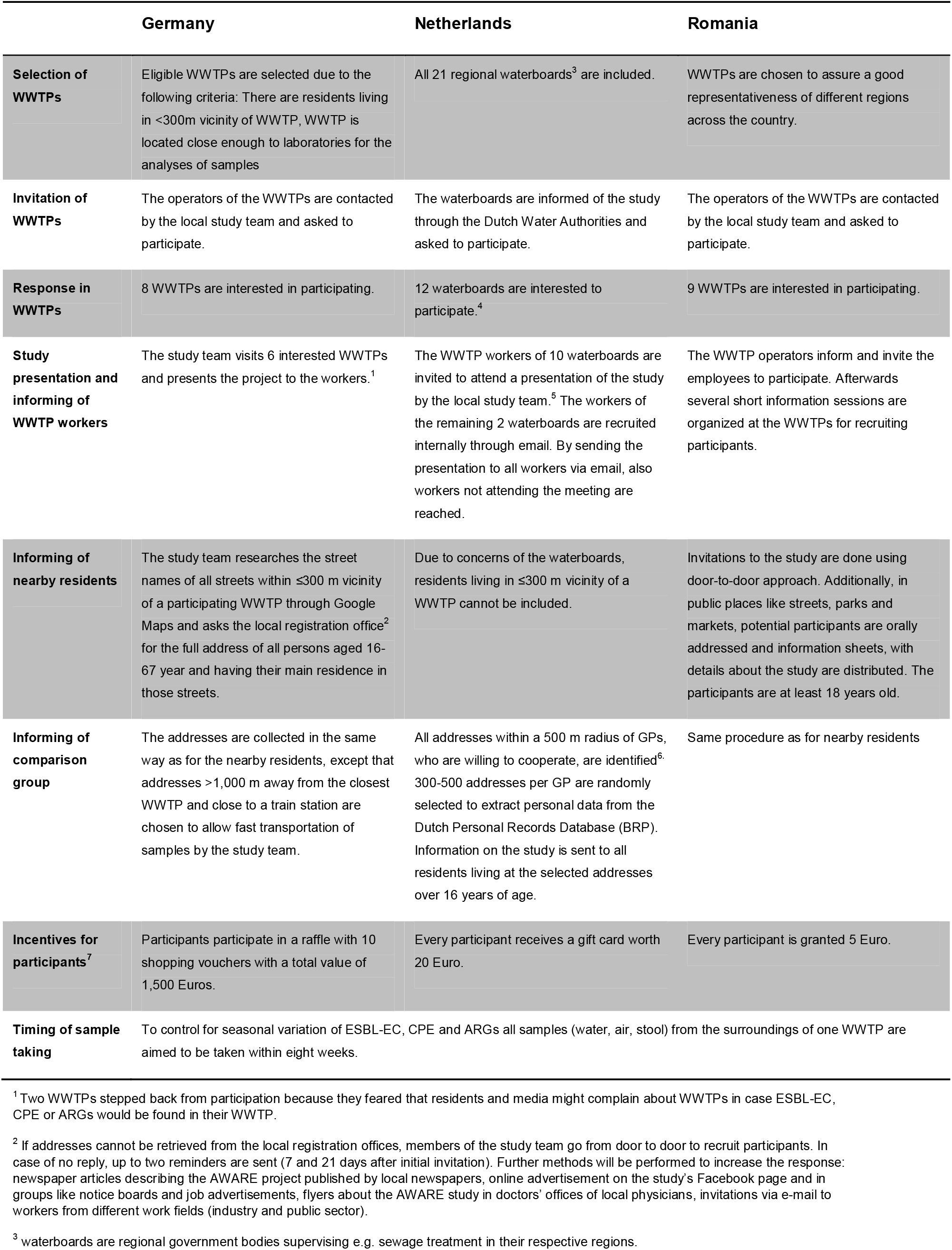
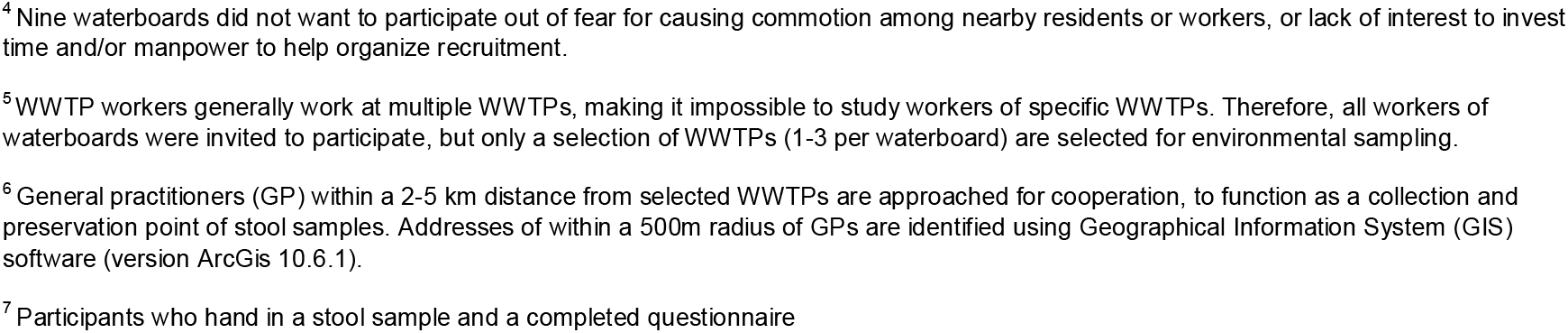
Recruitment of participants into the AWARE study.

|  | Germany | Netherlands | Romania |
| --- | --- | --- | --- |
| **Selection of WWTPs** | Eligible WWTPs are selected due to the following criteria: There are residents living in <300m vicinity of WWTP, WWTP is located close enough to laboratories for the analyses of samples | All 21 regional waterboards<sup>3</sup> are included. | WWTPs are chosen to assure a good representativeness of different regions across the country. |
| **Invitation of WWTPs** | The operators of the WWTPs are contacted by the local study team and asked to participate. | The waterboards are informed of the study through the Dutch Water Authorities and asked to participate. | The operators of the WWTPs are contacted by the local study team and asked to participate. |
| **Response in WWTPs** | 8 WWTPs are interested in participating. | 12 waterboards are interested to participate.<sup>4</sup> | 9 WWTPs are interested in participating. |
| **Study presentation and informing of WWTP workers** | The study team visits 6 interested WWTPs and presents the project to the workers.<sup>1</sup> | The WWTP workers of 10 waterboards are invited to attend a presentation of the study by the local study team.<sup>5</sup> The workers of the remaining 2 waterboards are recruited internally through email. By sending the presentation to all workers via email, also workers not attending the meeting are reached. | The WWTP operators inform and invite the employees to participate. Afterwards several short information sessions are organized at the WWTPs for recruiting participants. |
| **Informing of nearby residents** | The study team researches the street names of all streets within ≤300 m vicinity of a participating WWTP through Google Maps and asks the local registration office<sup>2</sup> for the full address of all persons aged 16-67 year and having their main residence in those streets. | Due to concerns of the waterboards, residents living in ≤300 m vicinity of a WWTP cannot be included. | Invitations to the study are done using door-to-door approach. Additionally, in public places like streets, parks and markets, potential participants are orally addressed and information sheets, with details about the study are distributed. The participants are at least 18 years old. |
| **Informing of comparison group** | The addresses are collected in the same way as for the nearby residents, except that addresses >1,000 m away from the closest WWTP and close to a train station are chosen to allow fast transportation of samples by the study team. | All addresses within a 500 m radius of GPs, who are willing to cooperate, are identified<sup>6</sup>. 300-500 addresses per GP are randomly selected to extract personal data from the Dutch Personal Records Database (BRP). Information on the study is sent to all residents living at the selected addresses over 16 years of age. | Same procedure as for nearby residents |
| **Incentives for participants<sup>7</sup>** | Participants participate in a raffle with 10 shopping vouchers with a total value of 1,500 Euros. | Every participant receives a gift card worth 20 Euro. | Every participant is granted 5 Euro. |
| **Timing of sample taking** | To control for seasonal variation of ESBL-EC, CPE and ARGs all samples (water, air, stool) from the surroundings of one WWTP are aimed to be taken within eight weeks. |  |  |

<sup>1</sup> Two WWTPs stepped back from participation because they feared that residents and media might complain about WWTPs in case ESBL-EC, CPE or ARGs would be found in their WWTP.
<sup>2</sup> If addresses cannot be retrieved from the local registration offices, members of the study team go from door to door to recruit participants. In case of no reply, up to two reminders are sent (7 and 21 days after initial invitation). Further methods will be performed to increase the response: newspaper articles describing the AWARE project published by local newspapers, online advertisement on the study's Facebook page and in groups like notice boards and job advertisements, flyers about the AWARE study in doctors' offices of local physicians, invitations via e-mail to workers from different work fields (industry and public sector).
<sup>3</sup> waterboards are regional government bodies supervising e.g. sewage treatment in their respective regions.
<sup>4</sup> Nine waterboards did not want to participate out of fear for causing commotion among nearby residents or workers, or lack of interest to invest time and/or manpower to help organize recruitment.
<sup>5</sup> WWTP workers generally work at multiple WWTPs, making it impossible to study workers of specific WWTPs. Therefore, all workers of waterboards were invited to participate, but only a selection of WWTPs (1-3 per waterboard) are selected for environmental sampling.
<sup>6</sup> General practitioners (GP) within a 2-5 km distance from selected WWTPs are approached for cooperation, to function as a collection and preservation point of stool samples. Addresses of within a 500m radius of GPs are identified using Geographical Information System (GIS) software (version ArcGis 10.6.1).
<sup>7</sup> Participants who hand in a stool sample and a completed questionnaire

### Pilot study

We test the study methods in a pilot phase which includes recruitment of study participants, the study questionnaire and sample taking (water, air, stool). The study questionnaire is tested by 33 participants. Fifteen participants hand in stool samples for the pilot study. Additionally, all six water and sludge samples are taken from two WWTPs.

### Study instruments

#### Study questionnaires

WWTP workers, residents and members of the comparison group willing to participate receive access to an online questionnaire. However, we offer paper questionnaires at the preference of the participants. For quality control, we do double data entry with error check. The questionnaire assesses socio-demographics (age, gender, education) as well as potential risk factors for ESBL, CPE or ARGs carriage (work history, travel abroad, contact with farm animals, hospital visits, antibiotic intake, self-evaluation of general health condition). Additionally, WWTP workers also answer questions considering their specific work tasks at the WWTP, the use of personal protective equipment and hygienic behaviour. WWTP operators answer questions about the capacity of their WWTP, origin of treated wastewaters, and wastewater treatment methods.

Whenever possible, we retrieve questions from validated questionnaires (Alavanja et al. 1996; Bisdorff et al. 2012; Burney et al. 1994; Council 1984; Heinrich et al. 2011; Nemeth 2006; O’Loughlin et al. 2015; Sandrock et al. 2007; Schutte et al. 2007a; Schutte et al. 2007b). Only if we cannot find validated questions, we take items from existing, but not validated questionnaires after checking for their face validity. If we cannot find any suitable questions from previous studies, we create expert validated new items. We translate the original questionnaires from English (supplemental 1, 2, 3) to German, Dutch and Romanian. At least two experts on the topic who are also native speakers of the target language check the translation and provide feedback. This pre-pilot phase of the study was an iterative process to translate, back-translate, ask for feedback and improve the current version of the questionnaires. We then test the translated questionnaires in a two-phase procedure: in the first phase, we recruit a small number of participants (n = 3) to read and provide verbal feedback on their understanding of each question. As we offer the questionnaire online, we create an online-survey using LimeSurvey (LimeSurvey GmbH). In the second phase, three persons of the target group go through the process of filling out the questionnaires online. They also provide feedback on the understanding of each question, and the online survey’s functionality. Once the questionnaire is refined and tested for clarity and understandability, it is tested in the pilot study. During the pilot study, seven WWTP operators (one from DE, six from RO) and twelve WWTP workers (three from DE, nine from RO), two nearby residents and twelve members of the comparison group fill in the questionnaire and provide feedback. Based on the results, we refine the questionnaire.

#### Stool samples

In DE and RO, participants receive a stool sample kit by postal service (residents and members of the comparison group) or at work (WWTP workers) after handing in an informed consent and completing the questionnaire. In NL, participants first hand in their stool samples and then fill in the questionnaire. We provide all necessary material to the participants in order to take the stool sample. This includes a paper faeces collection device, a sterile stool sampling tube and written and drawn instructions. In DE, participants are asked to bring the stool sample directly to the next WWTP, where it is cooled; or stored temporarily in a refrigerator until the next morning, when it is collected by a member of the study team. In NL, we ask participants working at a WWTP to bring their stool sample to the WWTP, where it is cooled, while residents are asked to bring it to a specified general practitioner (GP). GPs within a 2-5 km distance from selected WWTPs are approached for cooperation, to function as a collection and preservation point of stool samples. Addresses of within a 500m radius of GPs are identified using Geographical Information System (GIS) software (version ArcGis 10.6.1). Participants who are unable to bring their sample to the GP at the indicated time/day are given the opportunity to send the samples per mail without cooling (although samples shipped per mail will be excluded from metagenomic sequencing). In RO, we ask participants to cool the stool samples at 1-10°C directly after samples were taken and to bring them to the WWTP the next day. The same day, the stool samples are transported to the laboratory and processed within 72 hours. We tested this procedure in the pilot study with fifteen participants (one WWTP operator, three WWTP workers, and eleven members of the comparison group).

At the local laboratories in DE, NL and RO, all stool samples are inoculated directly onto the following agars: TBX or MacConkey, ChromID ESBL, ChromID OXA-48, ChromID CARBA and incubated at 36±2°C for 24 - 48h. In case of positive results, a total of two isolates belonging to the ESBL-EC phenotype and 5 isolates belonging to CPE phenotype are collected, screened for antibiotic resistance and identified by MALDI-TOF MS (*Matrix Assisted Laser Desorption Ionization-Time of Flight Mass Spectrometry*). We then process stool samples for DNA isolation after intermediate storage at -80°C, which we then will use for subsequent metagenomics and qPCR analyses.

#### Water samples

We collect water samples from WWTPs at four different treatment stages: wastewater influent (WI), effluent (WE), liquid sludge from the main biological reactor (e.g. aeriation tank) (AT), and dewatered sewage sludge after thickening (S). We also take water samples from the receiving surface water 200m upstream (WU) and 200m downstream (WD) of the WWTP. The following Figure 1 provides an overview of the collection points of water sample, as well as stool and air samples taken. We tested this procedure in the pilot study at one WWTP in DE and two in RO.

**Figure 1.**
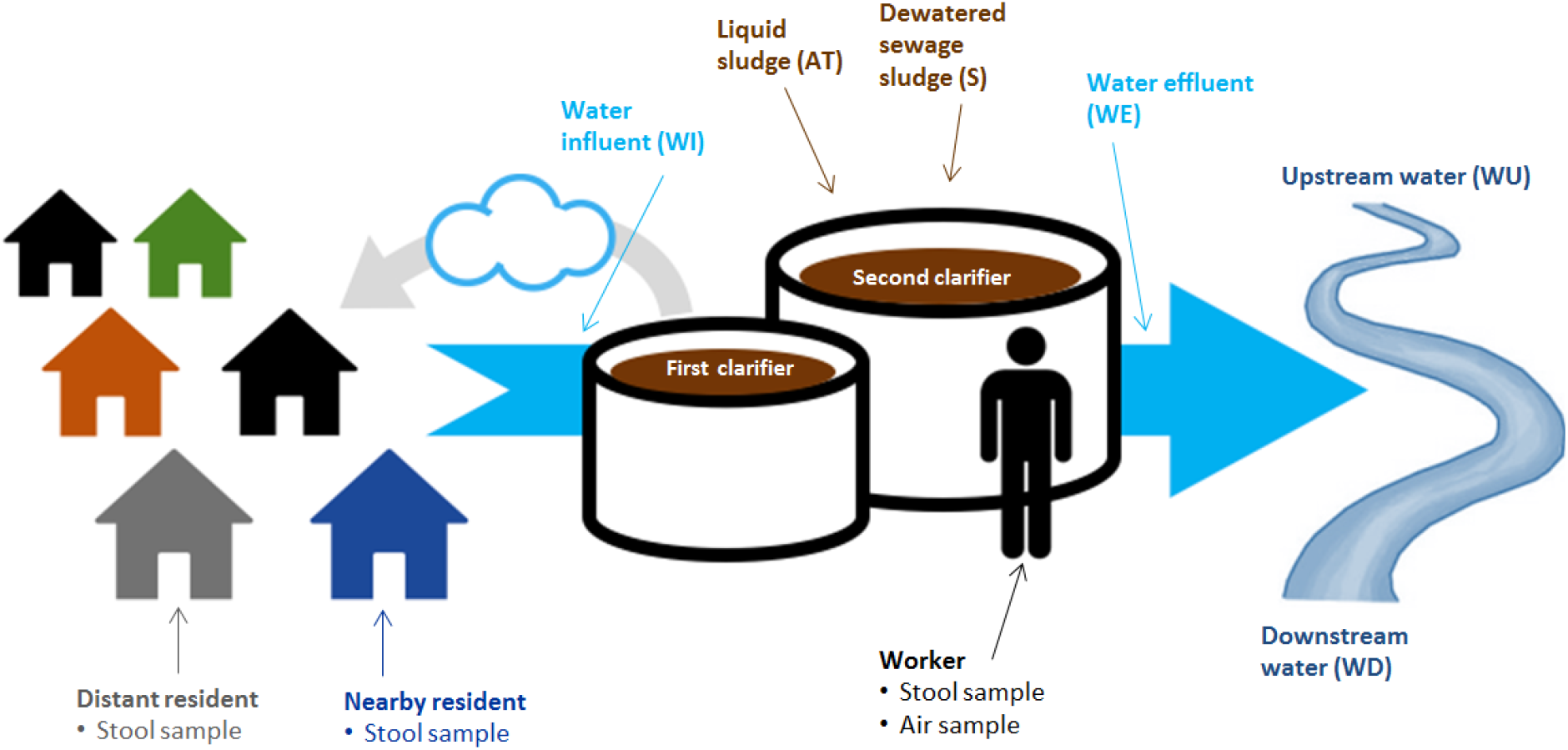
Collection points of water, air and stool samples.

We collect upstream (WU) and downstream (WD) water samples as close as possible to the WWTP to minimize the influence of other sources, but at enough distance to minimize the chance of diffusion to upstream locations and to ensure sufficient mixing with effluent for downstream locations. If accessible, we choose locations at 200m upstream and 200m downstream for waters with a width < 20m, according to the rule of thumb that complete mixing occurs at a distance of at least 10x the width of the surface water. Additionally, we choose the upstream and downstream locations in a way that no additional side streams enter the river between these locations and the effluent discharge point. Therefore, we choose locations closer to the WWTP when side streams are present within the optimal distance. We take subsurface samples according to international guidelines (ISO 19458:2007: Water quality - Sampling for microbiological analysis).

The sampling points for wastewater influent (WI) and effluent (WE) are determined by the location of the flow proportional auto samplers at the individual WWTPs, when present. Influent samplers are usually located directly after mechanical treatment and effluent samplers after completion of treatment, prior to discharge. Using auto samplers, experienced WWTP or laboratory staffs collect 24-h flow proportional samples, of which 1L is transferred to a sterile bottle at the end of the usual time interval applied in the WWTP (e.g., 9:00 in the morning). If no automatic samplers are available, we take grab samples from wastewater influent and effluent, at approximately 40 to 60 percent of the water depth, at a site with maximal turbulence to ensure good mixing and the possibility of solids settling is minimized. The most desirable sampling locations for grab samples of influent include: a) the upflow siphon following a comminutor (in absence of grit chamber); b) the upflow distribution box following pumping from main plant wet well; c) aerated grit chamber; d) flume throat; e) pump wet well when the pump is operating; or f) downstream of preliminary screening.

When possible, we take influent samples upstream from side stream returns. We collect grab samples of effluent at the site specified in the sampling plan, or if no site is specified, we select the most representative site downstream from all entering wastewater streams prior to discharge into the receiving waters.

We take the liquid sludge sample (AT) from the main biological reactor (e.g., aeration tank). The selection of the sampling points depends on a) the practicality of interrupting safely a stream of moving liquid sludge or cake when manually sampling; and b) the nature of the chamber or tank design with respect to stratification of liquid sludges.

We take the sample of dewatered sewage sludge after thickening (S). Prior to the proposed sampling date, we assess sludge processing (dewatering and treatment) to ensure that sludge is in the appropriate form (liquid versus dewatered, untreated cake versus treated biosolids), and is available for sampling at the proposed date, time, and sampling point. If needed, we will adjust the selection points.

After all water and sludge samples are collected, they are kept at 1-10°C at the WWTP and transported at 1-10°C to the laboratory in NL (samples from DE and NL) and RO (samples from RO). At the laboratories in NL and RO, we process all samples within 48-72 hours after sampling, e.g. homogenization (for sludge) and membrane filtration (for sludge and water). We then process water filters for DNA isolation, which we use for subsequent metagenomics and qPCR analyses.

#### Air samples

We intend to ask a subset of 50 workers from 10 WWTPs per participating country to collect air samples to analyse personal exposure. Sampling is based on GSP inhalable sampling heads equipped with Teflon filters on Gilair pumps (3.5 L/min), sampling the total inhalable air of workers whose job position included activities at different treatment stages.

The pumps are programmed and fixed at the worker’s belt or pocket by a member of the study team. A study team member checks the correct functioning of the pumps at the beginning, after three hours and after six hours of sampling. After six hours, the study team member turns off the pumps. We wrap the heads of the pumps in aluminium foil and transport them directly to the laboratory where the pumps are opened on a sterile work bench. The laboratory assistants remove the Teflon filters with a pair of sterile tweezers and freeze them at -20°C (DE) or -80°C respectively (NL, RO) in petri dishes. We ship all filters to NL for analysis. Feasibility of the procedures is checked during the pilot study.

### Metagenomic analysis

The Swedish and Romanian team conduct culture-independent analyses. They will employ shotgun metagenomics sequencing (Bengtsson-Palme et al. 2015; Bengtsson-Palme et al. 2014; Lundstrom et al. 2016) by the Illumina NovaSeq technology. This enables simultaneous quantification of any known antibiotic resistance gene if present at sufficiently high levels to allow detection. In addition, shotgun metagenomics allows for the analysis of mobile genetic elements such as integrons and transposons, and of the taxonomic composition of the microbial communities (Lundstrom et al. 2016). Although costs for DNA sequencing have dropped dramatically, it still involves substantial costs if relatively rare resistance genes are targeted in complex community samples (Bengtsson-Palme et al. 2015; Gweon et al. 2019). Therefore, we will select a subset of air, sewage, water, and faecal samples for sequencing, while we plan to choose 12 genes for qPCR investigations in all human, water and air samples. The selection will be based on an initial screen using qPCR arrays with considerably more genes for a subset of samples. Antibiotic residues and their metabolites are usually detected in the environment at trace levels, but may still be present at concentrations that have the potential to select for microbial resistance (Bengtsson-Palme and Larsson 2016; Lundstrom et al. 2016) and possibly also induce horizontal gene transfer (Jutkina et al. 2018). Therefore, residues are monitored by high-performance liquid chromatography interfaced with tandem mass spectrometry (HPLC–MS/MS) in selected plants, including the WWTPs in which metagenomics data are also determined. We perform sample selection for metagenomic analyses by using propensity score matching of the exposed and unexposed groups to achieve proportional and non-statistically significant balance of the groups at a 5% statistical level.

### Data management

We store the personal contact data of participants and the history of contacts via letters, e-mails and phone calls in a password protected Access database separated from questionnaire and sample data. We pseudonymize all assessed data. The laboratories document results of stool, air, and water samples in Excel. We primarily do data cleaning and analysis in R. Additional software will be used depending on the specific analyses. All personal data are stored password protected with access only to the members of the study team. We ensure that data management is bound to FAIR principles (Wilkinson et al. 2016; Wilkinson et al. 2019), e.g. including storage of research data obtained in publicly accessible and findable repositories.

### Statistical analysis

For descriptive analyses we assess the distribution of numerical variables visually for normality using histograms, and present the mean ± standard deviation if normally distributed or the median ± inter-quartile range if non-normally distributed. We present categorical variables using absolute and relative frequencies. We handle missing values by multiple imputation in case of missing at random or missing completely at random. We do data cleaning, as well as multiple imputation, propensity score matching, data presentation, and outcome models using the statistical software R version 3.5 and up (R-Core-Team 2019). Additional software will be used and documented depending on the specific analyses.

We perform bivariate hypothesis testing choosing an appropriate statistical test depending on the type of variables involved, their distribution and the number of counts per cell (for categorical variables). We perform logistic crude and adjusted regression models for the main outcomes such as carriage of ESBL-EC, CPE and ARGs. Main exposure variables will include whether a participant belongs to the group of WWTP workers, nearby residents or the comparison group. We consider linear regression models for secondary outcomes if these are numerical. We present results from regression models with the point estimate and its corresponding 95% confidence interval. We do variable selection for the models using a combination of experts’ opinion from within the AWARE consortium, evidence in the current literature, and the use of Directed Acyclic Graphs (DAGs).

### Ethics and informed consent

Ethics approval was received by the responsible ethics committees of the participating study centres in DE (Committee: Ethikkommission bei der medizinsichen Fakultăt der LMU München, number of approval: 17-734) and RO (Committee: Comisia de Etică a Cercetării, number of approval: 164/05.12.2017). In NL this research is exempted for ethical approval under the Dutch Medical Research Involving Human Subjects Act (WMO; Committee: Medisch Ethische Toetsingscommissie, number of confirmation: 19-001/C).

All work adheres to established conventions governing such research, in particular, Directive 95/46/EC, the Helsinki declaration, and the 1977 Oviedo Convention of the Council of Europe on human rights and biomedicine. A signed informed consent is retrieved from all study participants.

### Funding Statement

AWARE (Antibiotic Resistance in Wastewater: Transmission Risks for Employees and Residents around Wastewater Treatment Plants) is supported by the European Commission (JPI-EC-AMR ERA-Net Cofund grant no 681055), in Germany with the “Bundesministerum für Bildung und Forschung” with DLR Projektträger (grant 01KI1708), in the Netherlands with JPI AMR, ZonMw (grant 547001007), in Romania with UEFISCDI project ERANET-JPI-EC-AMR-AWARE-WWTP (grant 26/2017) and in Sweden with Swedish Research Council VR (grant 2016-06512).

## Discussion

To our knowledge, this is the first study investigating the potential spreading of ESBL-EC, CPE and ARGs from WWTP to workers, the environment and nearby residents. By involving different European countries, covering a variety of different types of WWTPs, our results will be relevant for a large number of situations. The methodological combination of epidemiology, molecular biology and metagenomics will allow us to draw multilevel conclusions. We demonstrated feasibility of the AWARE project in the pilot study.

Our study is carried out cross-sectionally at each WWTP. Thus, the study does not provide information how the numbers of ESBL-EC, CPE and ARGs varies with time/seasons. It is possible that bias arises for some samples due to different laboratories analysing them. In order to minimize such biases, we develop all SOPs jointly and centralize sample preparation and analyses whenever possible. WWTP workers are organized in different ways depending on the country: In NL, WWTP workers do not work at one specific WWTP, hampering the comparison between ESBL-EC, CPE and ARGs at the selected WWTP and in stool from workers.

Our assessment of transmission of antibiotic resistant bacteria from WWTPs to the surrounding environment will enable us to formulate recommendations, such as adapted sewage treatment in order to reduce antibiotic resistant bacteria or recommendations for a minimal distance between WWTPs and residential buildings.

## Data Availability

This publication describes the methods of a cross-sectional study. Since the study is not finalized yet, data sets are not finalized and are therefore are not available yet.

## Notes

### Competing Interest Statement

The authors have declared no competing interest.

### Clinical Trial

This publication describes the methods of a cross-sectional study for which a trial ID is not necessary.

### Author Declarations

Ethics approval was received by the responsible ethics committees of the participating study centres in DE (Committee: Ethikkommission bei der medizinsichen Fakultaet der LMU Muenchen, number of approval: 17-734) and RO (Committee: Comisia de Etică a Cercetării, number of approval: 164/05.12.2017). In NL this research is exempted for ethical approval under the Dutch Medical Research Involving Human Subjects Act (WMO; Committee: Medisch Ethische Toetsingscommissie, number of confirmation: 19-001/C).

