## Supplemental 1 for "Antibiotic Resistance in Wastewater Treatment Plants and Transmission Risks for Employees and Residents: The Concept of the AWARE Study"

KLINIKUM
DER UNIVERSITÄT MÜNCHEN

CAMPUS CITY CENTRE

INSTITUTE AND OUTPATIENT CLINIC FOR OCCUPATIONAL; SOCIAL AND ENVIRONMENTAL MEDICINE

### Questionnaire

#### Resistance in Wastewater: Transmission Risks for Employees and Residents around Wastewater

OCCUPATIONAL AND ENVIRONMENTAL EPIDEMIOLOGY & NET TEACHING UNI

Director: PROF. DR. KATJA RADON, MSC
Director: PROF. DR. MED. DENNIS NOWAK

AWARE

Project coordinators:
Dr. Laura Wengenroth, Dr. Tobias Weinmann
Institute and Outpatient Clinic for Occupational, Social and Environmental Medicine
Clinical Centre of the Ludwig-Maximilian-University (LMU)
Ziemssenstr. 1, 80336 München
Telefon: 089/4400-52483
Telefax: 089/4400-54954

Study-ID:

Dear participant,

thank you for your interest in participating in the AWARE study. In this questionnaire we will ask you several questions about you, your work and family environment and your health. Please answer as completely and accurately as possible. If you are unsure about a question, please select “no”. Filling out the questionnaire should take you around 20 minutes.

Thank you very much!

Your AWARE-Team

CONTACT
Dr. Laura Wengenroth, Dr. Tobias Weinmann

Website: www.aware-study.eu

Klinikum der Universität München
Ziemssenstr. 1
80336 München

Postanschrift: Postfach 15 20 25 · 80051 München

**Directors**

**Prof. Dr. Katja Radon** · Institute and Outpatient Clinic for Occupational, Social and Environmental Medicine, Clinical Centre of the Ludwig-Maximilian University (LMU) Munich, Germany

**Prof. Dr. Ana Maria de Roda Husman** · National Institute for Public Health and the Environment (RIVM), Bilthoven, Niederlande

**Prof. Dr. Carmen Chifiriuc** · Research Institute of the University of Bucharest (ICUB), Bukarest, Rumänien

**Prof. Dr. Joakim Larsson** · Institute of Biomedicine, University of Gothenborg, Göteborg, Schweden

Please follow the following instructions to help you fill out the questionnaire:

Please mark your answer to each question by marking the area inside the answer box with a cross as shown in the example.

EXAMPLE: ☒

If you make a mistake and need to correct it, please fill in the whole box.

EXAMPLE: ■

To answer open questions, please write clearly and in block letters on the corresponding row. If a number is required, please write it clearly in the corresponding field.

EXAMPLE: 16

Please make sure to answer all questions by going line by line. You may skip questions only when the text explicitly indicates it..

EXAMPLE: No....... ☒ -> Please continue with question XY
ja .........☐ 1

If you mark "yes", please continue on with the next question. If you mark "no", please proceed only to the question indicated by the arrow.

Please chack again for completeness after you have answered the questionnaire.

**Please do not forget to fill in the <u>informed consent</u>!**
**We will not be able to evaluate your questionnaire without your signed consent!**

#### SOCIO-DEMOGRAPHICS

1. **What is your date of birth?**

| __ | __ |  | __ | __ | __ | __ |
   Month  Year

2. **In which country were you born?**

   ______________________________________

3. **Are you male of female?**
   ☐ male
   ☐ female

4. **What is your education level? If you have several degrees, please choose the highest!**
   ☐ Primary education
   ☐ Lower secondary education
   ☐ Degree of a technical college
   ☐ Higher secondary education
   ☐ Other graduation
   ☐ Left school without education
   ☐ Not yet graduated

#### JOB HISTORY

***Please answer "yes" or "no" whether possible. If you are unsure, please answer with "no".***

5. **Have you worked in the past 12 months?**
   Every occupation you have had for at least 1 months is relevant. It does not matter whether you worked at home (e.g. househusband/housewife) or out of home, full-time or part-time, paid or unpaid or self-employed (e.g. in a family business).
   ☐ No → Please continue with question 12
   ☐ Yes

6. **Which kind of jobs and /or internships have you had <u>within the last 12 months</u>?**
   If you have more than one job in the same company or have two different jobs at the same time, please name them separately. Please begin with your most recent occupation.

| Occupation/ Job | Line of Work | When did you start working in this job? (month/year) | If applicable, when did you stop working in this job? (month/year) | Hours per week |
|---|---|---|---|---|
| | | I__I__I/I__I__I__I__I | I__I__I/I__I__I__I__I | I__I__I |
| | | I__I__I/I__I__I__I__I | I__I__I/I__I__I__I__I | I__I__I |
| | | I__I__I/I__I__I__I__I | I__I__I/I__I__I__I__I | I__I__I |
| | | I__I__I/I__I__I__I__I | I__I__I/I__I__I__I__I | I__I__I |
| | | I__I__I/I__I__I__I__I | I__I__I/I__I__I__I__I | I__I__I |

7. **Have you worked in a livestock farm <u>in the last 12 months</u>?**
   ☐ No
   ☐ Yes

8. **Have you been in contact with animal manure at work <u>in the past 12 months</u>?**
   If you are unsure, please answer with "No"!
   ☐ No
   ☐ Yes

9. **<u>Within the past 12 months</u>, have you personally performed slaughterhouse work of slaughterhouse activities?**
   ☐ No
   ☐ Yes

10. **In your current job, how often have you typically had direct interaction or contact with patients <u>within the last 12 months</u>?**
    ☐ Never ☐ Rarely ☐ Sometimes ☐ Often ☐ Always

11. **How often have you worked with human tissue, blood, body fluids (urine, faeces, vomit, sputum, saliva) or primary cell lines <u>within the last 12 months</u>?**
    ☐ Never ☐ Rarely ☐ Sometimes ☐ Often ☐ Always

#### ENVIRONMENT

***Please answer "yes" or "no" wherever possible. If you are unsure, please answer with "no".***

**12. Have you travelled abroad <u>in the past 12 months</u>?**

☐ No → Please continue with question 15
☐ Yes

**13. How many times have you travelled to the following continents <u>in the past 12 months</u>?**

| | Never | Once | 2-3 times | More than 3 times | Don't know |
|---|---|---|---|---|---|
| Europe | ☐ | ☐ | ☐ | ☐ | ☐ |
| North Africa (north of the Sahara) | ☐ | ☐ | ☐ | ☐ | ☐ |
| Africa (except North Africa) | ☐ | ☐ | ☐ | ☐ | ☐ |
| Asia | ☐ | ☐ | ☐ | ☐ | ☐ |
| North America | ☐ | ☐ | ☐ | ☐ | ☐ |
| Central America / Mexico | ☐ | ☐ | ☐ | ☐ | ☐ |
| South America | ☐ | ☐ | ☐ | ☐ | ☐ |
| Australia and Oceania | ☐ | ☐ | ☐ | ☐ | ☐ |

**14. If you have travelled to Europe, have you been in one of the following countries: Italy, Slovenia, Bulgaria or Greece?**

☐ No
☐ Yes

**15. Do you currently live in a farm?**

☐ No
☐ Yes

**16. Is your home located within <u>500 m</u> from one or more animal stalls?**

☐ No
☐ Yes, from one
☐ Yes, from two
☐ Yes, from more than two, in fact |_|_| stalls (please enter the number)

**17. Is your workplace located within <u>500 m</u> from one or more animal stalls?**

☐ No
☐ Yes, from one
☐ Yes, from two
☐ Yes, from more than two, in fact |_|_| stalls (please enter the number)

**18. Have you regularly visited agricultural facilities keeping animals (at least once a month) <u>within the last 12 months</u> (e.g. to buy milk or eggs)?**

☐ No → Please continue with question 20
☐ Yes

**19. If yes, how often?**

☐ less than once per week
☐ 1-3 days per week
☐ 4-7 days per week

**20. <u>In the last 12 months</u> have you entered the barn of the following animals?**

| | No | Yes | Don't know |
|---|---|---|---|
| Pigs | ☐ | ☐ | ☐ |
| Poultry | ☐ | ☐ | ☐ |
| Dairy cattle, calves | ☐ | ☐ | ☐ |

**21. How often <u>in the last 12 months</u> have you entered the following barns?**

| | Less than once per week | 1-3 days per week | 4-7 days per week |
|---|---|---|---|
| Barn for pigs | ☐ | ☐ | ☐ |
| Barn for poultry | ☐ | ☐ | ☐ |
| Barn for dairy cattle or calves | ☐ | ☐ | ☐ |

**22. Did you own any of the following animals <u>in the last 12 months</u>?**

| | No | Yes |
|---|---|---|
| Horse | ☐ | ☐ |
| Dog | ☐ | ☐ |
| Cat | ☐ | ☐ |

#### HOSPITAL VISITS AND MEDICATION

***Please answer "yes" or "no" wherever possible. If you are unsure, please answer with "no".***

**23. <u>In the last 12 months</u>, have you been in a hospital for <u>more than 12 hours</u>?**

☐ No → Please continue with question 26
☐ Yes

**24. Why were you i the hospital for more than 12 hours? (Multiple answers possible)**

☐ Visitor → Please continue with questions 26
☐ Professional → Please continue with question 26
☐ Patient

**25. Did you have any surgery?**

☐ No
☐ Yes

**26. Do you have any chronic condition that requires frequent (at least once a month) hospital visits? (e.g. dialysis kidney dysfunction, cancer, others….)**
☐ No
☐ Yes → **How frequent?** |_|_| times per week *or* |_|_| times per month

**27. Have you taken an antibiotic <u>within the last 12 months</u>?**
☐ No
☐ Yes
☐ Do not know

**<u>Antibiotic:</u>** A drug for bacterial infectious diseases

**28. In the last 12 months, have you used or taken any antacids? (Heartburn remedies, gastric protection, gastric acid neutralization agents, proton pump inhibitors, e.g. omeprazole, esomeprazole, lansoprazole, dexlansoprazole, rabeprazole)**
☐ No
☐ Yes
☐ Do not know

**<u>Antacid:</u>** An agent that neutralizes acidity or reduces production, especially in the digestive system.

#### YOUR HEALTH AND WELL BEING

***Please answer "yes" or "no" wherever possible. If you are unsure, please answer with "no".***

**29. <u>In the last 12 months</u>, how often did you have loose, mushy or watery stools?**
☐ Never ☐ Rarely ☐ Sometimes ☐ Often ☐ Always

**30. In general, would you say your health is**
☐ Excellent ☐ Very good ☐ Good ☐ Fair ☐ Poor

**31. How much do you feel bothered by any odors in your living environment coming from the neighbourhood (especially from the wastewater treatment plant)?**
☐ Not at all ☐ A little ☐ Clearly ☐ Strongly

***The next few questions will be mostly about your breathing. Please answer "yes" or "n" wherever possible. If you are unsure, please answer with "no".***

**32. Have you ever had wheezing or whistling in your chest at any time <u>in the last 12 months?</u>**
☐ No → Please continue with question 34
☐ Yes

**33. Have you had this wheezing or whistling when you <u>did not</u> have a cold?**
☐ No
☐ Yes

**34. Have you had an attack of shortness of breath that came on during the day when you were at rest at any time in the last 12 months?**
☐ No

☐ Yes

**35. Have you had an attack of shortness of breath that came on FOLLOWING strenuous activity in the last 12 months?**
☐ No
☐ Yes

**36. Have you ever been woken up by an attack of shortness of breath at any time in the last 12 months?**
☐ No
☐ Yes

**37. Do you cough on most days for as much as 3 months each year?**
☐ No → Please continue with question 39
☐ Yes

**38. If yes, have you had this affliction for at least 2 years?**
☐ No
☐ Yes

**39. Do you bring up any phlegm on most days for as much as 3 months each year?**
☐ No → Please continue with questions 41
☐ Yes

**40. If yes, have you had this affliction for at least 2 years?**
☐ No
☐ Yes

**41. Have you ever had asthma?**
☐ No → Please continue with questions 43
☐ Yes

**42. Was this confirmed by a doctor?**
☐ No
☐ Yes

#### ABOUT YOUR FAMILY

***Please answer "yes" or "no" wherever possible. If you are unsure, please answer with "no".***

**43. Do you have children? (Including adopted children or stepchildren)**
☐ No → Please continue with question 45)
☐ Yes

**44. How many children do you have? (Including adopted children or stepchildren)**
|__|__| children

**45. Besides yourself, are there other people living in your household?**
☐ No → Please continue with question 51
☐ Yes
☐ Prefer not to say → Please continue with question 51

**46. Please indicate which of the following people (besides you) live in your household**

| | No | Yes | How many |
|---|---|---|---|
| Mother | ☐ | ☐ | \|_\|_\| |
| Father | ☐ | ☐ | \|_\|_\| |
| Partner/Spouse | ☐ | ☐ | \|_\|_\| |
| Children | ☐ | ☐ | \|_\|_\| |
| Others, Who (e.g. grandmother)? | | | |
| • ____________________ | ☐ | ☐ | \|_\|_\| |
| • ____________________ | ☐ | ☐ | \|_\|_\| |
| • ____________________ | ☐ | ☐ | \|_\|_\| |
| • ____________________ | ☐ | ☐ | \|_\|_\| |

**47. Is a member of your household (besides you) currently working in one of the following areas? (multiple answers possible**

☐ Farm
☐ Slaughterhouse
☐ Nursing Home
☐ Physician's practice or hospital
☐ None of the mentioned above

**48. How many times has a member of your household (besides you) travelled to the following continents in the past 12 months?**

| | Never | Once | 2-3 times | More than 3 times | Don't know |
|---|---|---|---|---|---|
| **Europe** | ☐ | ☐ | ☐ | ☐ | ☐ |
| **North Africa (North of the Sahara)** | ☐ | ☐ | ☐ | ☐ | ☐ |
| **Africa (except North Africa)** | ☐ | ☐ | ☐ | ☐ | ☐ |
| **Asia** | ☐ | ☐ | ☐ | ☐ | ☐ |
| **North America** | ☐ | ☐ | ☐ | ☐ | ☐ |
| **Central America/Mexico** | ☐ | ☐ | ☐ | ☐ | ☐ |
| **South America** | ☐ | ☐ | ☐ | ☐ | ☐ |
| **Australia and Oceania** | ☐ | ☐ | ☐ | ☐ | ☐ |

**49. Has a member of your household (besides you) been hospitalized for more than 12 hours in the last 12 months?**

☐ No
☐ Yes

**50. Have any of your family members (besides you) taken antibiotics <u>within the last 12 months</u>?**

☐ No
☐ Yes
☐ Do not know

**<u>Antibiotic:</u>** A drug for bacterial infectious diseases

#### *<u>Questions for WWTP employees</u>*

#### CURRENT WORK IN WWTPS

***All these questions refer to the wastewater treatment plant in which you received this questionnaire or the link to the questionnaire.***

**51. Through which wastewater treatment plant did you receive information about this study?**

________________________________________

**52. Do you work in another wastewater treatment plant besides the one where you received the questionnaire?**

☐ No → Please continue with question 55
☐ Yes

**53. In which other wastewater treatment plant(s) have you worked?**

Wastewater treatment plant 1: _________________________
Wastewater treatment plant 2: _________________________
Wastewater treatment plant 3: _________________________
Wastewater treatment plant 4: _________________________
Wastewater treatment plant 5: _________________________
Wastewater treatment plant 6: _________________________
Wastewater treatment plant 7: _________________________
Wastewater treatment plant 8: _________________________
Wastewater treatment plant 9: _________________________
Wastewater treatment plant 10: _________________________

**54. Since when have you worked in these wastewater treatment plants?**

| | **Month** | **Year** |
|---|---|---|
| Wastewater treatment plant 1: | I__I__I | I__I__I__I__I |
| Wastewater treatment plant 2: | I__I__I | I__I__I__I__I |
| Wastewater treatment plant 3: | I__I__I | I__I__I__I__I |
| Wastewater treatment plant 4: | I__I__I | I__I__I__I__I |
| Wastewater treatment plant 5: | I__I__I | I__I__I__I__I |
| Wastewater treatment plant 6: | I__I__I | I__I__I__I__I |
| Wastewater treatment plant 7: | I__I__I | I__I__I__I__I |

| | Month | Year |
|---|---|---|
| Wastewater treatment plant 8: | |__|__| | |__|__|__|__|
| Wastewater treatment plant 9: | |__|__| | |__|__|__|__|
| Wastewater treatment plant 10: | |__|__| | |__|__|__|__|

**55. Do you work in the following wastewater treatment plant facilities? If so, how often do you work here?**
***If steps are combined, please answer "yes" if you are exposed to each sub step.***

| | Never | Less than once per week | 1-3 days per week | 4-7 days per week |
|---|---|---|---|---|
| Air release value or manhole | ▪☐ | ▪☐ | ☐ | ☐ |
| Pumping station | ▪☐ | ▪☐ | ☐ | ☐ |
| Headworks | ▪☐ | ▪☐ | ☐ | ☐ |
| Screening | ▪☐ | ▪☐ | ☐ | ☐ |
| Pre-aeration | ▪☐ | ▪☐ | ☐ | ☐ |
| Grit removal | ▪☐ | ▪☐ | ☐ | ☐ |
| Flow equalization | ☐ | ☐ | ☐ | ☐ |
| Septage receiving and handling facilities | ▪☐ | ▪☐ | ☐ | ☐ |
| Side stream returns (including digester decant, dewatering return flows, or backwash water) | ☐ | ☐ | ▪ ☐ | ▪ ☐ |
| Primary clarifier | ▪☐ | ▪☐ | ▪☐ | ▪☐ |
| Aeration basin | ▪☐ | ▪☐ | ▪☐ | ▪☐ |
| Secondary clarifier | ▪☐ | ▪☐ | ▪☐ | ▪☐ |
| Return sludge | ▪☐ | ▪☐ | ▪☐ | ▪☐ |

**56. Do you work in the following sludge facilities? If so, how often do you work here?**

| | Never | Less than once a week | 1-3 days per week | 4-7 days per week |
|---|---|---|---|---|
| Sludge thickening tank | ☐ | ☐ | ☐ | ☐ |
| Wet sludge storage | ☐ | ☐ | ☐ | ☐ |
| Wet sludge loadout facilities | ☐ | ☐ | ☐ | ☐ |
| Digestion | ☐ | ☐ | ☐ | ☐ |

| | Never | Less than once a week | 1-3 days per week | 4-7 days per week |
|---|---|---|---|---|
| Mechanical dewatering | ☐ | ☐ | ☐ | ☐ |
| Sludge storage after mechanical dewatering | ☐ | ☐ | ☐ | ☐ |
| Dry sludge loadout facilities | ☐ | ☐ | ☐ | ☐ |
| Composting facilities | ☐ | ☐ | ☐ | ☐ |
| Alkaline stabilisation (stabilised solids generation resulting from reaction with lime) | ☐ | ☐ | ☐ | ☐ |
| Sludge drying bed | ☐ | ☐ | ☐ | ☐ |

**57. Do you perform cleaning operations at work?**

☐ No → Please continue with question 59
☐ Yes

**58. During cleaning work, do you use a high-pressure cleaner? If yes, how often?**

☐ No
☐ Yes: ___ hours/day *or* _____ days/week

**59. Do you use treated effluent for cleaning?**

☐Never ☐Rarely ☐Sometimes ☐Often ☐Always

**60. Do you eat at other places besides offices or canteens during work?**

☐Never ☐Rarely ☐Sometimes ☐Often ☐Always

**61. If yes, where do you eat?** _________________________

#### PROTECTIVE MEASURES

**62. Do you use protective clothing or equipment while working?**

☐ No
☐ Yes, but just for specific tasks
☐ Yes, most of the time at work

**63. How often do you use protective gloves or breathing protection?**

**64. After your workday, do you take a shower?**

| : | Everyday | Several times per week | Several times per month | Several times per year | Not at all | For which asks? |
|---|---|---|---|---|---|---|
| Protective gloves | ☐ | ☐ | ☐ | ☐ | ☐ | |
| Surgical mask | ☐ | ☐ | ☐ | ☐ | ☐ | |
| Face mask including filter | ☐ | ☐ | ☐ | ☐ | ☐ | |
| Face mask with air pressure | ☐ | ☐ | ☐ | ☐ | ☐ | |
| Other (which one): ______________ | ☐ | ☐ | ☐ | ☐ | ☐ | |
| Other (which one): ______________ | ☐ | ☐ | ☐ | ☐ | ☐ | |

☐ No → Please continue with question 66
☐ Yes, at work → Please continue with question 66
☐ Yes, at home → Please continue with question 66
☐ No, generally not, only after specific activities

**65. After which specific activities do you take a shower?** ________________________

**66. How often do you change your work clothes?**
☐ Never → Please continue with question 68
☐ Once a month → Please continue with question 68
☐ Once a week → Please continue with question 68
☐ After specific activities

**67. Please list those activities:** ________________________

**68. Where do you wash your working clothes?**
☐ At home
☐ At work
☐ A company washes my clothes

**69. Do you put on your work clothes at home or at work?**
☐ At home
☐ At work

**Thank you very much for participating in the AWARE Study!**

With your help we came closer to achieving our goals of the study

If you have any further questions, please do not hesitate to contact us via our webpage www.aware-study.eu or via.

Here, you have the possibility to make comments:

______________________________________________________________________
______________________________________________________________________
______________________________________________________________________
______________________________________________________________________
______________________________________________________________________
______________________________________________________________________

**Have you already signed the informed consent? Otherwise, we cannot include your questionnaire data in the AWARE Study!**

AWARE

Project coordinators:
Dr. Laura Wengenroth, Dr. Tobias Weinmann
Institute and Outpatient Clinic for Occupational, Social and Environmental Medicine
Clinical Centre of the Ludwig Maximilian University (LMU)
Ziemssenstr. 1, 80336 München
Telephone: 089/4400-52483
Telefax: 089/4400-54954
