## Supplemental 2 for "Antibiotic Resistance in Wastewater Treatment Plants and Transmission Risks for Employees and Residents: The Concept of the AWARE Study"

LMU KLINIKUM DER UNIVERSITÄT MÜNCHEN

CAMPUS INNENSTADT

INSTITUT UND POLIKLINIK FÜR ARBEITS-, SOZIAL- U. UMWELTMEDIZIN

### Questionnaire

#### Antibiotic Resistance in Wastewater: Transmission Risks for Employees and Residents around Wastewater Treatment Plants

OCCUPATIONAL AND ENVIRONMENTAL EPIDEMIOLOGY & NET-TEACHING UNIT

Head: PROF. DR. KATJA RADON, MSC

AWARE

Study team: Dr. Laura Wengenroth, Dr. Tobias Weinmann
Institute and Outpatient Clinic for Occupational, Social and Environmental Medicine
Clinical Centre of the Ludwig Maximilian University (LMU)
Ziemssenstr. 1, 80336 München
Telephone: 089/4400-52483
Telefax: 089/4400-54954


Study-ID:

Dear participant,

thank you for your interest in participating in the AWARE study. In this questionnaire we will ask you several questions about you, your work and family environment and your health. Please answer as completely and accurately as possible. If you are unsure about a question, please select "no". Filling out the questionnaire should take you around 20 minutes.

Thank you very much!

Your AWARE-Team

**Contact**
Dr. Laura Wengenroth, Dr. Tobias Weinmann

Website: www.aware-study.eu

Klinikum der Universität München
Ziemssenstr. 1
80336 München

Postal address: Postfach 15 20 25 · 80051 München

**Study Coordination**

**Prof. Dr. Katja Radon** · Institute and Outpatient Clinic for Occupational, Social and Environmental Medicine, Clinical Centre of the Ludwig Maximilian University (LMU) Munich, Germany

**Prof. Dr. Ana Maria de Roda Husman** · National Institute for Public Health and the Environment (RIVM), Bilthoven, Netherlands

**Prof. Dr. Carmen Chifiriuc** · Research Institute of the University of Bucharest (ICUB), Bucharest, Romania

**Prof. Dr. Joakim Larsson** · Institute of Biomedicine, University of Gothenborg, Göteborg, Sweden

#### Please follow the following instructions to help you fill out the questionnaire:

Please mark your answer to each question by marking the area inside the answer box with a cross as shown in the example:

Example: 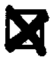

If you make a mistake and need to correct it, please fill in the complete box:

Example: 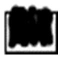

To answer open questions, please write clearly and in block letters on the corresponding row. If a number is required, please write it clearly in the corresponding field:

Example: 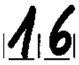

years

Please make sure to answer all questions by going line by line. You may skip questions only when the text explicitly indicates it.

No 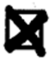 ............. → Please continue with question XY

Yes ............ ☐

If you mark „yes“, please continue on with the next question. If you mark „no“, please proceed only to the question indicated by the arrow!

Please check again for completeness after you have answered the questionnaire.

### GENERAL INFORMATION ON WWTP

1. **Is the amount of inhabitants served known?**
   ☐ No → Please continue with question 3
   ☐ Yes

2. **If yes, how many inhabitants does the wastewater treatment plant serve?**
   I__I__I__I__I__I__I__I inhabitants

3. **What is the typical daily flow (dry weather conditions)?**
   I__I__I__I__I__I__I $m^3$/day

4. **What is the maximal daily flow (wet weather conditions)?**
   I__I__I__I__I__I__I $m^3$/hour

5. **What is the design capacity in population equivalents?**
   ____________________p.e.
   ***Population equivalent (p.e.)*** *is a term used to measure the organic biodegradable load generated in an urban area. It takes into account the load generated by the resident population, the non-resident population (e.g. tourists) and industries. A population equivalent of 1 is definfed as the organic biodegradable load having a fiveday biochemical oxygen demand of 60 g of oxygen per day or 120 g COD/day*
   *COD = chemical oxygen demand*

6. **What is the actual utilization (in population equivalents p.e.)?**
   ____________________p.e.

7. **What is the food to microorganism ratio (F/M-ratio) in g BOD/kg d.s./day?**
   ____________________ g BOD/kg d.s./day
   ***BOD*** *= biochemical oxygen demand*
   *d.s. = dry solids*

8. **On which biological method is the treatment based?**
   ☐ Activated sludge process → Please continue with question 10
   ☐ Others

9. **On which other biological methods is the treatment based?**

   ________________________________________________________________________

10. **Does this WWTP treat hospital wastewater?**
    ☐ No → Please continue with question 81
    ☐ Yes
    ☐ Do not know → Please continue with question 81

11. **If yes, from how many hospitals?**
    I__I__I hospitals

12. **Does this WWTP treat wastewater from nursing homes / elderly peoples' homes?**

☐ No → Please continue with question 14
☐ Yes
☐ Do not know → Please continue with question 14

**13. If yes, from how many nursing homes / elderly peoples' homes?**

|__|__|__|

**14. Is the following treatment existing in the plant: <u>Primary sedimentation</u>?**

☐ No
☐ Yes

**15. Is the following treatment existing in the plant: <u>Aeration, biological stages</u>?**

☐ No → Please continue with question 17
☐ Yes

**16. How many biological stages exist?**

☐ One stage
☐ Two stages

**17. Is the following treatment existing in the plant: <u>Nitrification</u>?**

☐ No
☐ Yes

**18. Is the following treatment existing in the plant: <u>Denitrification</u>?**

☐ No
☐ Yes

**19. Is the following treatment existing in the plant: <u>Phosphorus removal</u>?**

☐ No → Please continue with question 21
☐ Yes

**20. Does phosphorous removal take place chemically or biologically?**

☐ Chemically
☐ Biologically
☐ Chemically and biologically

**21. Is the following treatment existing in the plant: <u>Solid separation</u>?**

☐ No
☐ Yes

**22. Is the following treatment existing in the plant: <u>Advanced treatment such as disinfection of effluent or removal of micropollutants</u>?**

☐ No → Please continue with question 24
☐ Yes

**23. Which advanced treatment for desinfection of effluent or removal of micropollutants do you use?**

______________________________

24. **Is the following treatment existing in the plant: <u>Open sludge storage</u>?**
   ☐ No
   ☐ Yes

25. **Sludge treatment is present for the following streams:**
   ☐ Sludge thickening
   ☐ Sludge digestion
   ☐ Sludge dewatering

26. **Are the following wastewater treatment facilities present at the WWTP: <u>Air release valve or manhole</u>?**
   ☐ No
   ☐ Yes

27. **Are the following wastewater treatment facilities present at the WWTP: <u>Pumping station</u>?**
   ☐ No → Please continue with question 29
   ☐ Yes

   28. **Is it a wet well or dry well pumping station?**
      ☐ Wet well
      ☐ Dry well

29. **Are the following wastewater treatment facilities present at the WWTP: <u>Septage receiving and handling facilities</u>?**
   ☐ No
   ☐ Yes

30. **Are the following wastewater treatment facilities present at the WWTP: <u>Side stream returns (including digester decant, dewatering return flows, or backwash water)</u>?**
   ☐ No
   ☐ Yes

31. **Are the following wastewater treatment facilities present at the WWTP: <u>Return sludge</u>?**
   ☐ No → Please continue with question 33
   ☐ Yes

   32. **How is return sludge transported?**
      ☐ Pump
      ☐ Headworks

33. **Are the following wastewater treatment facilities present at the WWTP? Are these open or closed: <u>Headworks</u>?**
   *"Open" means that residents may experience odor nuisance, while "closed" means that residents can not experience any odor nuisance.*

   ☐ No

☐ Yes, open
☐ Yes, enclosed

**34. Are the following wastewater treatment facilities present at the WWTP? Are these open or closed: <u>Screenning</u>?**
☐ No
☐ Yes, open
☐ Yes, enclosed

**35. Are the following wastewater treatment facilities present at the WWTP? Are these open or closed: <u>Pre-aeration</u>?**
☐ No
☐ Yes, open
☐ Yes, enclosed

**36. Are the following wastewater treatment facilities present at the WWTP? Are these open or closed: <u>Grit removal</u>?**
☐ No
☐ Yes, open
☐ Yes, enclosed

**37. Are the following wastewater treatment facilities present at the WWTP? Are these open or closed: <u>Flow equalization</u>?**
☐ No
☐ Yes, open
☐ Yes, enclosed

**38. Are the following wastewater treatment facilities present at the WWTP? Are these open or closed: <u>Primary Clarifier</u>?**
☐ No
☐ Yes, open
☐ Yes, enclosed

**39. Are the following wastewater treatment facilities present at the WWTP? Are these open or closed: <u>Aeration Basin</u>?**
☐ No → Please continue with question 42
☐ Yes, open
☐ Yes, enclosed

**40. How does wastewater treatment take place in the aeration basin?**
☐ Anoxic
☐ Aerobic
☐ Anaerobic

**41. How does aeration take place in the aeration basin?**
☐ Cascade
☐ Vertical axis surface aerator
☐ Fine bubble aeration
☐ Frush aeration
☐ Other

42. **Are the following wastewater treatment facilities present at the WWTP? Are these open or closed: <u>Trickling filter</u>?**
☐ No
☐ Yes, open
☐ Yes, enclosed

43. **Are the following wastewater treatment facilities present at the WWTP? Are these open or closed: <u>Secondary Clarifier</u>?**
☐ No
☐ Yes, open
☐ Yes, enclosed

44. **Are the following sludge and biosolids facilities present at the WWTP? Are these open or closed?**

*"Open" means that residents may experience odor nuisance, while "closed" means that residents can not experience any odor nuisance.*

| | **No** | **Yes, open** | **Yes, enclosed** |
|---|---|---|---|
| Sludge thickening | ☐ | ☐ | ☐ |
| Sludge storage after mechanical dewatering | ☐ | ☐ | ☐ |
| Composting facilities | ☐ | ☐ | ☐ |
| Sludge drying bed | ☐ | ☐ | ☐ |

45. **Are the following sludge and biosolids facilities present at the WWTP: <u>Wet sludge storage</u>?**
☐ No → Please continue with question 47
☐ Yes

46. **Is the wet sludge storage covered or uncovered?**
☐ Covered
☐ Uncovered

47. **Are the following sludge and biosolids facilities present at the WWTP: <u>Wet sludge loadout facilities</u>?**
☐ No → Please continue with question 49
☐ Yes

48. **Are the wet sludge ladaout facilities covered or uncovered?**
☐ Covered
☐ Uncovered

49. **Are the following sludge and biosolids facilities present at the WWTP: <u>Digestion</u>?**
☐ No → Please continue with question 51
☐ Yes

**50. How does digestion take place?**

☐ Aerobic
☐ Anaerobic, mesophilic
☐ Anaerobic, thermophilic

**51. Are the following sludge and biosolids facilities present at the WWTP: <u>Mechanical Dewatering</u>?**

☐ No → Please continue with question 53
☐ Yes

**52. How does mechanical dewatering take place?**

☐ Belt filter press
☐ Recessed plate filter press
☐ Centrifuge
☐ Other

**53. Are the following sludge and biosolids facilities present at the WWTP: <u>Dry sludge loadout facilities</u>?**

☐ No → Please continue with question 55
☐ Yes

**54. Are the dry sludge loadout facilities covered or uncovered?**

☐ Covered
☐ Uncovered

**55. Are the following sludge and biosolids facilities present at the WWTP: <u>Alkaline stabilization (stabalized solids generation resulting from reaction with lime)</u>?**

☐ No
☐ Yes

#### Thank you very much for participating in the AWARE Study!

With your help we came closer to achieving our goals of the study.

If you have any further questions, please do not hesitate to contact us via our webpage www.aware-study.eu or via.

Here, you have the possibility to make comments:

AWARE

Dr. Laura Wengenroth, Dr. Tobias Weinmann
Institute and Outpatient Clinic for Occupational, Social and Environmental Medicine
Clinical Centre of the Ludwig Maximilian University (LMU)
Ziemssenstr. 1, 80336 München
Telephone: 089/4400-52483
Telefax: 089/4400-54954
